# Does the elevated (thrombosis risk of males relative to females help account for the excess male mortality observed in Covid-19? An observational study

**DOI:** 10.1101/2021.05.04.21256001

**Authors:** Kenneth Cohen, David Anderson, Sheng Ren, David J. Cook

## Abstract

**Objectives:** We sought to determine whether underlying thrombophilia helps account for excess Covid-19 mortality rates in males relative to females. Specifically, we asked: What is the contribution of thrombophilia to the excess Covid-19 mortality risk among malesã

**Design:** Cross-sectional observational study.

**Setting:** Data were sourced from electronic medical records (EMRs) drawn from over 200 US hospital systems.

**Participants:** 16,576 patients hospitalized with Covid-19, aged 40 and above.

**Main outcome measures:** The primary study outcome was Covid-19 mortality. We measured: 1) the mortality rate of male patients relative to female patients, 2) the rate of thrombotic diagnoses occurring during hospitalization for Covid-19 in male and female patients, and 3) the mortality rate when evidence of thrombosis was present. We used hospital EMR data to determine the rates of a thrombotic diagnosis by sex, and D-dimer levels to help identify undiagnosed thrombosis.

**Results:** The Covid-19 mortality rate of males was higher than that of females by 16.1%. Males with Covid-19 also had a 25.4% higher rate of receiving a thrombotic diagnosis compared to females with Covid-19. The mortality rate of all patients with a thrombotic diagnosis was 42.4%—a rate over twice that of Covid-19 patients without a thrombotic diagnosis (adjusted OR 2.4 (2.17 to 2.65), p-value < .001). When defining thrombosis as either a documented thrombotic diagnosis or markedly elevated D-dimer levels, over half of the excess mortality in male patients could be explained by thrombophilia.

**Conclusions:** Our findings suggest the higher Covid-19 mortality rate in males may be significantly accounted for by the increased propensity for thrombophilia among males. This thrombotic tendency is additive to the thrombotic risk associated with Covid-19. Understanding the mechanisms that underlie male thrombophilia may allow for the advancement of effective anticoagulation strategies that reduce Covid-19 mortality.

**Strengths and limitations of this study:**

- To our knowledge, this population study is the first to provide evidence of a sex-based clinical association that may help explain the excess Covid-19 mortality in males compared to females.
- This was an observational study using diagnosis codes and natural language processing (NLP) extractions of detailed electronic health record (EHR) data for 16,576 hospitalized patients with Covid-19.
- We evaluated underlying thrombosis based on both an inpatient thrombotic diagnosis as well as a markedly elevated D-dimer level relative to CRP level.
- A limitation of this study was that a portion of the estimated excess male mortality was based on the analysis of a subset of 3,442 patients that had serial D-dimer and CRP levels measured in the hospital, and that a potential undercount of these patients was possible due to the use of NLP.

## Introduction

Male sex has emerged as a risk factor for increased COVID-19 morbidity and mortality, with the risk of mortality in males being 30%-40% higher than in females.^1, 2^ This heightened mortality risk for males is independent of advancing age, and cannot be fully explained by higher rates among males of other co-morbidities known to increase mortality in COVID-19, such as obesity, diabetes mellitus, hypertension, or underlying cardiopulmonary disease.^3, 4^ Therefore, the etiology of this sex difference in COVID-19 mortality is largely unexplained.

We also know that thrombosis appears to play an important role in the morbidity and mortality of COVID-19, with patients at increased risk of both micro- and macrovascular thrombosis. In a recent meta-analysis of 42 studies representing data from 8,271 COVID-19 patients, researchers discovered an overall venous thromboembolism rate of 21% and a pulmonary embolism rate of 13%. For patients admitted to the ICU, the rate of thromboembolism was even higher, at 31%. Notably, the pooled odds of mortality were 74% higher among COVID-19 patients who developed thromboembolism, compared to those who did not.^5^

The thrombophilia of males relative to females was well-established in the medical literature long before the emergence of COVID-19. In multiple studies of unprovoked deep venous thrombosis and pulmonary embolus (DVT/PE) where hereditary thrombophilia was excluded, the rate of recurrent thrombosis in males has been reported to be anywhere from two to more than three times higher than females.^6-10^ Despite extensive research on the topic, the etiology of this sex-based difference in male thrombophilia remains unknown.^11^

We therefore understand that males are at risk for higher Covid-19 mortality, that COVID-19 mortality risk is increased when patients have underlying thrombosis, and that unrelated to COVID-19, males are more prone to thrombosis relative to females. It is therefore plausible that the inherent thrombophilia in males relative to females may partially explain the excess mortality rate observed in males with COVID-19. In this study we sought to understand whether males with COVID-19 have a higher rate of thrombosis compared to females with COVID-19, and whether a higher rate of thrombosis contributes to the excess mortality seen in males.

## Methods

### Data Source

To explore a potential connection between thrombosis, sex, and mortality risk, we analyzed hospitalization data from over 200 geographically dispersed hospital systems. From this data we selected patients aged 40 and above who were hospitalized for Covid-19 between March 3 and October 22, 2020. We limited our analysis to those patients who survived and were discharged from the hospital, those discharged to hospice care, those who died in the hospital, and those who died after being discharged to home. Patients who died soon after discharge to home or who were discharged to hospice care were treated as deceased cases due to Covid-19. Covid-19 readmission cases were excluded from our study.

The data for this study was sourced from EMR data and post-EMR coding. These EMRs were processed with a natural language processing (NLP) engine that produces a homogenized set of codified and non-codified information, including lab results, medications, symptoms, and various observational extracts of text from the EMR. The at-scale extraction of this detailed information allows insight into the clinical manifestations of the Covid-19 population. We examined the subsets of patients who survived Covid-19 compared to those who expired. For each of the groups, we compared the male and female incidence of receiving a thrombosis diagnosis code while hospitalized for Covid-19.

### Outcomes and Study Variables

The primary outcome of interest was mortality from Covid-19. The exposure variable was biological sex, and the key variable of interest was thrombosis. We classified thrombotic diagnosis codes into four major conditions: (1) myocardial infarction (MI), (2) DVT/PE, (3) stroke, and (4) peripheral artery occlusion (PAO). A Full list of ICD-10 codes used to identify thrombosis can be found in Supplementary Table 1.

Because thrombotic diagnoses are underreported in inpatients with Covid-19,^12^ we also examined the peak D-dimer value of the subset of hospitalized patients in whom this was measured. Markedly elevated D-dimer, out of proportion to other acute phase reactants such as CRP levels, has been associated with increased mortality in Covid-19, presumably due to underlying thrombosis.^13^ For the purpose of quantifying the proportion of excess mortality attributed to a thrombophilia, we used D-dimer as a surrogate measure for a thrombotic diagnosis when a thrombotic diagnosis code was absent but the D-dimer level was markedly elevated.

### Statistical Methods

To determine whether the inherent thrombophilia in males is associated with higher Covid-19 mortality, we used mediation analysis,^14^ where thrombophilia was the mediator between the outcome (mortality) and the exposure variable (gender). Mediation analysis consists of three steps, which are illustrated in Figure 1. First, we needed to confirm that male patients had higher mortality rate than female patients in our study population. Second, we needed to confirm that males had a higher incidence of a thrombosis diagnosis relative to females. Third, we needed to demonstrate that both males and females with thrombosis diagnoses had a higher mortality rate than those who did not. If all three of these conditions are supported by statistically significant differences between groups, we can combine regression models from the first and third hypothesis to estimate the proportion of sex effect on mortality that can be explained by thrombophilia. In estimating this proportion, we examined the effect of a thrombosis diagnosis together with evidence of markedly elevated D-dimer levels, recognizing that such levels are also likely indicative of underlying thrombosis.

**Figure 1.** Mediation analysis flowchart.

Logistic regression models were used in each of the three steps. The response variables and covariates of interest are illustrated in Figure 1. We used the Wald test for regression coefficients in logistic regression models to assess statistical significance in all steps. All hypothesis tests were one-sided at the 0.05 significance level. We report 95% confidence intervals and one-tailed p-values. All regression models were adjusted for age and co-morbidities derived from the Charlson Index.^15^

The proportion of excess male mortality explained by thrombophilia is defined by the ratio of the mediation effect to the total effect (mediation + direct effect) of gender on mortality. This proportion was estimated using R packages “mediation”^16^ and “mma”.^17^ Because thrombotic diagnoses are underreported in inpatients as discussed above, and because the association between D-dimer values and mortality could be non-linear, we estimated the proportion in several different ways. First, we based our estimate on the presence of a thrombosis diagnoses only. Next, we used an expanded thrombophilia definition that included D-dimer values > 2.0 µg/mL^18^ along with the presence of a thrombotic diagnosis. Finally, we used the expanded thrombophilia definition along with actual D-dimer values, assuming linear, piecewise linear (using D-dimer values = 2.0 µg/mL as threshold), and non-linear relationships to mortality. To remove the possibility that elevated D-dimer levels represented an acute phase reactant, we also adjusted C-reactive protein (CRP) values in regression models. This adjustment allowed us to attribute the proportion of mortality explained by a thrombosis diagnosis and D-dimer values to thrombophilia rather than to severe acute phase reactions. The upper bounds of D-dimer and CRP normal ranges (0.5 µg/mL and 10 mg/L respectively) were used to normalize D-dimer and CRP values.

### Patient and Public Involvement

As this study represents an exploratory analysis of de-identified patient data, neither patients nor the public were involved in the design, conduct, reporting, or dissemination plans of our research.

## Results

Our study population was comprised of 16,576 Covid-19 patients with an average age of 65.1 years. There were slightly more male patients (53.1%) than female patients. Among study patients, 16.8% had a reported thrombotic diagnosis during their hospital stay and 23.0% died or were discharged to hospice care. We also identified a subset of 3,442 patients (20.8%) out of the study population without a thrombotic diagnosis who had complete peak D-dimer and CRP values. Normalized mean peak D-dimer and CRP values during a hospital stay were 5.26 (526% of the normal range upper bound) and 4.51 (451% of the normal range upper bound). Characteristics of the full study population and this patient subset are shown in Table 1.

**Table 1.** Characteristics of the Full Hospitalized Population and the D-dimer Subset

| Study sample characteristics | Summary statistics<sup>a</sup> |  |
| --- | --- | --- |
|  | Full population dataset | D-dimer analysis subset |
| Sample Size | 16,576 | 3,442 |
| Deceased | 3,814 (23.01) | 773 (22.46) |
| Thrombophilia | 2,786 (16.81) | 525 (15.25) |
| Thrombosis History | 1,145 (6.91) | 194 (5.64) |
| Mechanical Ventilation | 2,550 (15.38) | 625 (18.16) |
| Deceased or Intubated | 4,534 (27.35) | 964 (28.01) |
| MI | 1,029 (6.21) | 165 (4.79) |
| DVT | 1,418 (8.55) | 298 (8.66) |
| Stroke | 568 (3.43) | 102 (2.96) |
| PAO | 84 (0.51) | 10 (0.29) |
| Age average | 65.07 | 64.54 |
| Sex: male | 8,798 (53.08) | 1,843 (53.54) |
| D-dimer average | ----- | 525.72% |
| CRP average | ----- | 451.20% |
| D-dimer median | ----- | 266% |
| CRP median | ----- | 169% |

Abbreviations: MI, myocardial infarction; DVT, deep vein thrombosis; PAO, peripheral arterial occlusion; CRP, C-reactive protein.
<sup>a</sup> With the exception of D-dimer and CRP statistics, sample size, and age, data are otherwise reported as number (percentage) of patients within each dataset. D-dimer and CRP statistics are based on the upper bound of the normal range for each measure.

We found that the mortality rate in males with Covid-19 was higher by 16.4% compared to females with Covid-19 (an absolute rate difference of 3.4%; adjusted odds ratio (OR) = 1.38 (1.27 to 1.50), p < .001). Compared to females with Covid-19, males with Covid-19 had a rate of receiving a thrombotic diagnosis during their hospital stay that was 25.4% higher (an absolute difference of 3.8%, OR = 1.29 (1.19 to 1.41), p < .001), confirming the higher rate of thrombotic diagnoses in males. Additionally, we found an over two-fold difference in mortality between patients with and without a thrombotic diagnosis (42.4% vs. 19.1%; adjusted OR = 2.4 (2.17 to 2.65), p < .001). Having verified that thrombophilia is more prevalent in males (OR = 1.29) and that thrombosis is a strong risk factor for mortality (OR = 2.4), we conclude that thrombophilia accounts for a portion of the excess mortality in males.

Based on these findings, we then sought to determine what proportion of the excess male mortality in Covid-19 patients might be related to thrombophilia. When using only a documented diagnosis of thrombosis, the proportion of the mortality effect explained by thrombosis was significant at 11% (p < .001) (Table 2). Because of underreporting of thrombotic diagnoses in hospitalized patients with Covid-19, we also considered a markedly elevated D-dimer level out of proportion to an elevated CRP level as a surrogate of underlying thrombosis. We therefore combined the proportion of increased mortality in males as predicted by a thrombotic diagnosis with the proportion explained by the D-dimer analysis. In doing so, the total proportion of excess male mortality potentially explained by thrombophilia increased to 22.9% (9.22% to 45.4%) when the thrombophilia definition was expanded to include D-dimer levels > 2.0 µg/mL, and up to 53.6% (21.7% to 87.9%) when D-dimer values were treated as continuous measures in nonlinear regression models (Table 3).

**Table 2.** Full Mediation Analysis Results of Thrombosis Diagnosis

| Mediation analysis steps |  |  |  |
| --- | --- | --- | --- |
| **Step 1:**<br>**Documentation of the higher mortality rate in males** |  | **Female** | **Male** |
|  | Number of patients | 7,778 | 8,798 |
|  | Survived | 6,129 | 6,633 |
|  | Deceased | 1,649 | 2,165 |
|  | Sex distribution among deceased patients | 43.2% | 56.8% |
|  | Rate of deceased patients by sex | 21.2% | 24.6% |
|  | Unadjusted odds ratio | 1.21 (1.13, 1.3) p-value < .001 |  |
|  | Adjusted odds ratio | 1.38 (1.27, 1.5) p-value < .001 |  |
| **Step 2:**<br>**Documentation of the higher rate of thrombosis in males** |  | **Female** | **Male** |
|  | Number of patients | 7,778 | 8,798 |
|  | No thrombosis diagnosis | 6,626 | 7,164 |
|  | Had thrombosis diagnosis | 1,152 | 1,634 |
|  | Sex distribution among thrombosis patients | 41.4% | 58.7% |
|  | Thrombosis prevalence by sex | 14.8% | 18.6% |
|  | Unadjusted odds ratio | 1.31 (1.21, 1.42) p-value < .001 |  |
|  | Adjusted odds ratio | 1.29 (1.19, 1.41) p-value < .001 |  |
| **Step 3:**<br>**Documentation of the higher mortality rate associated with a thrombosis diagnosis** |  | **No thrombosis** | **Had thrombosis** |
|  | Number of patients | 13,790 | 2,786 |
|  | Survived | 11,156 | 1,606 |
|  | Deceased | 2,634 | 1,180 |
|  | Thrombosis distribution among deceased patients | 69.1% | 30.9% |
|  | Mortality rates by thrombosis | 19.1% | 42.4% |
|  | Unadjusted odds ratio | 3.11 (2.85, 3.39) p-value < .001 |  |
|  | Adjusted odds ratio | 2.40 (2.17, 2.65) p-value < .001 |  |
|  | Adjusted odds ratio for sex | 1.34 (1.23, 1.46) p-value < .001 |  |
|  | Proportion Mediated (Explained) | 11.0% (6.08%, 17.4%) p-value < .001 |  |

**Table 3.** Proportion of Excess Mortality Explained by the Addition of D-dimer Values to the Expanded Thrombophilia Diagnosis

| Mediators | Model | Proportion mediated |
| --- | --- | --- |
| Thrombosis diagnosis codes or D-dimer > 2.0 µg/mL | Linear | 22.9% (9.22%, 45.4%) |
| Thrombosis diagnosis codes or D-dimer > 2.0 µg/mL + D-dimer value | Linear | 25.7% (11.9%, 48.4%) |
| Thrombosis diagnosis codes or d-dimer > 2.0 µg/mL + D-dimer value + D-dimer–CRP ratio | Piecewise Linear<sup>b</sup> | 36.1% (20.6%, 62.0%) |
| Thrombosis diagnosis codes or D-dimer > 2.0 µg/mL + D-dimer value | Non-linear | 53.6% (21.7%, 87.9%) |

<sup>b</sup> A D-dimer value of 2.0 µg/mL was used to assign different regression coefficients to the D-dimer value and the D-dimer–CRP ratio, based on whether the D-dimer value was above or below this threshold.

## Discussion

### Statement of principle findings

The data extracted from our large population of hospitalized patients with Covid-19 confirms a higher mortality rate among those patients with thrombosis. It also demonstrates that the incidence of thrombosis is higher in males relative to females. When multiple indicators of thrombophilia are considered, it appears that thrombophilia in males may explain over half of the excess mortality seen in males with Covid-19.

### Strengths and weaknesses in relation to other studies

To our knowledge, this is the first study to specifically address the significantly elevated risk of thrombosis and its associated excess mortality in males with Covid-19 relative to females. Although mortality as a function of sex was not specifically addressed in the meta-analysis of the 42 studies noted in the introduction,^5^ that study also documented that thrombosis contributes to excess mortality in COVID-19. Moreover, among those 42 studies, 29 documented the sex of the patients. Of those 29 studies, 27 (93%) documented a higher percentage of thromboembolism in males relative to females. Of all cases of thromboembolism across those 29 studies, 70% occurred in males and 30% occurred in females.

Studies of prophylactic treatment for thrombosis among Covid-19 patients also suggest that greater attention to thrombosis risk may be warranted. A recent systematic review and pooled analysis of 35 studies looked at anticoagulation strategies in 4,685 hospitalized patients with Covid-19.^19^ This review showed standard prophylactic doses of anticoagulation were associated with significant reductions in venous thromboembolism and arterial thrombosis events, with intermediate and therapeutic doses of anticoagulation providing no additional benefit. Data from these studies did not allow for an analysis of dose intensity by sex.

### Unanswered questions and future research

Because the known genetic risks for thrombophilia (factor V Leiden, prothrombin G20210A, protein C and S abnormalities, etc.) are not sex-linked chromosomal mutations, they cannot explain this excess male risk. There are, however, genetic mutations found on the X and Y chromosomes which might explain a portion of this increased thrombophilia in males.^11^ Additionally, severe Covid-19 infection is itself associated with a prothrombotic state, the mechanisms of which are not fully understood. Possible mechanisms for activation of the coagulation cascade include direct viral invasion of the vascular endothelium, activation as part of the generalized cytokine activation seen in severe Covid-19, and the development of procoagulant autoantibodies, including antiphospholipid antibodies.

### Conclusion

While there has been extensive research on male thrombophilia prior to the emergence of Covid-19, the mechanism of this increased male thrombotic tendency remains speculative, as do the mechanisms that underlie the thrombotic risk intrinsic to Covid-19. Understanding the mechanisms that drive increased male thrombophilia as well as those that drive thrombosis in Covid-19 may allow for the development of more effective anticoagulation strategies that reduce the mortality risk for both males and females diagnosed with Covid-19. To help develop these strategies, randomized clinical trials are needed to test more intense anticoagulation regimens in males hospitalized with Covid-19.

## Data Availability

Data availability statement: The data are not available for public use but, under certain conditions, may be made available to editors and their approved reviewers under a data-use agreement to confirm the findings of the current study.

## Author Contributions

Dr. Ken Cohen, Mr. Dave Anderson, Dr. Sheng Ren, and Dr. David Cook had full access to all of the data in the study and take responsibility for the integrity of the data and the accuracy of the data analysis.

### Concept and design

All authors.

### Acquisition, analysis, or interpretation of data

All authors.

### Drafting of the manuscript

All authors.

### Critical revision of the manuscript for important intellectual content

All authors and Amy Okaya, MPH, MBA, OptumLabs.

### Editing of manuscript

Amy Okaya.

### Statistical analysis

Sheng Ren.

### Supervision

Ken Cohen.

## Competing interests

All authors have completed the ICMJE uniform disclosure form at http://icmje.org/disclosure-of-interest/ and declare: all authors had financial support from UnitedHealth Group for the submitted work; no financial relationships with any organisations that might have an interest in the submitted work in the previous three years; no other relationships or activities that could appear to have influenced the submitted work.

## Funding

The study was funded by OptumLabs, the research and development arm of UnitedHealth Group, and the authors Drs. Cohen, Cook, Ren, and Mr. Anderson are full-time employees of UnitedHealth Group. These authors played an active role in all aspects of study development, including the design and conduct of the study; collection, management, analysis, and interpretation of the data; preparation, review, or approval of the manuscript; and decision to submit the manuscript for publication.

## Ethics approval

Because no identifiable protected health information was extracted or accessed during the course of the study and all data were accessed in compliance with the Health Insurance Portability and Accountability Act, Institutional Review Board approval, other ethics approval, or waiver of authorization was not required.

## Transparency statement

The lead author* affirms that this manuscript is an honest, accurate, and transparent account of the study being reported; that no important aspects of the study have been omitted; and that any discrepancies from the study as planned (and, if relevant, registered) have been explained.

*The manuscript’s guarantor

## Data availability statement

The data are not available for public use but, under certain conditions, may be made available to editors and their approved reviewers under a data-use agreement to confirm the findings of the current study.

## Notes

### Competing Interest Statement

The authors have declared no competing interest.

### Author Declarations

This manuscript was provided an IRB waiver as it was a retrospective analysis with all patient level data blinded. This was provided by the Office of Human Research Affairs at United Health Group, Emily Robison, Senior Regulatory Consultant

