## Supplementary material for "Does the elevated (thrombosis risk of males relative to females help account for the excess male mortality observed in Covid-19? An observational study": Figure 1

### Mediation Analysis Flowchart

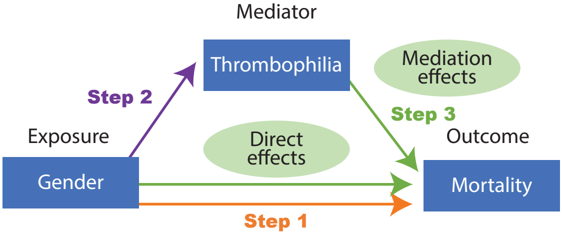


Each arrow (step) represents a regression model pointing to the response variable. The origin of each arrow represents the covariate of interest of the response variable.
